# Evaluation of lymphocyte subtypes in COVID-19 patients

**DOI:** 10.1101/2021.07.12.21260382

**Authors:** Mitra Rezaei, Majid Marjani, Payam Tabarsi, Afshin Moniri, Mihan purabdollah, Zahra Abtahian, Mehdi Kazempour Dizaji, Neda Dalil Roofchayee, Neda K. Dezfuli, Davood Mansouri, Nikoo Hossein-Khannazer, Mohamad Varahram, Esmaeil Mortaz, Ali Akbar Velayati

## Abstract

**Background:** Although the many aspects of COVID-19 have not been yet recognized, it seems that the dysregulation of the immune system has a very important role in the progression of the disease. In this study the lymphocyte subsets were evaluated in COVID-19 patients with different severity.

**Methods:** In this prospective study, the levels of peripheral lymphocyte subsets (CD3^+^, CD4^+^, CD8^+^ T cells; CD19^+^ and CD20^+^ B cells; CD16^+^/CD56^+^ NK cells, and CD4^+^/CD25^+^/FOXP3^+^ regulatory T cells) were measured in 67 confirmed patients with COVID-19 on the first day of admission.

**Results:** The mean age of cases was 51.3 ± 14.8 years. Thirty-two patients (47.8%) were classified as severe cases and 11 (16.4%) patients were categorized as critical. The frequency of blood lymphocytes, CD3^+^ cells, CD25^+^FOXP3^+^ T cells; and absolute count of CD3^+^ T cells, CD25^+^FOXP3^+^ T cells, CD4^+^ T cells, CD8^+^ T cells, CD16^+^56^+^ lymphocytes were lower in more severe cases in comparison to milder cases. Percentages of lymphocytes, T cells, and NK cells were significantly lower inthe patients who died (p= 0.002 and P= 0.042, p=0.006, respectively).

**Conclusion:** Findings of this cohort study suggests that the frequency of CD4^+^, CD8^+^, CD25^+^FOXP3^+^ T cells, and NK cells were difference in the severe COVID-19 patients. Moreover, lower frequency of, T cells, and NK cells are predictors of mortality of these patients.

## Introduction

Coronavirus disease 19 (COVID-19) was first reported from Wuhan, China in December 2019. The etiologic agent was an emerging coronavirus named severe acute respiratory syndrome coronavirus 2 (SARS-CoV-2). In a short period, COVID-19 spread to other countries led to a pandemic [1].Globally, by 17 May 2021, there have been 163 M confirmed cases of COVID-19, including 3.3M deaths, reported to the world health organization [2]. The first report from Iran reported on the 19^th^ of February 2020 and by 25 October 2020, 574,856 confirmed cases, and 32,953 deaths [3]. Although about 80 percent of affected patients have mild symptoms, the disease can be severe enough to need admission among 20 % of patients, and 5% led to respiratory or other organ failure and even leads to death [4].

Despite the global effort to determine the pathophysiology of COVID-19, the many aspects of the disease have not been recognized to date. It seems that the dysregulation of the immune system has a very important role in the progression of COVID-19 [5, 6]. Lymphocytes and the subsets especially cytotoxic T lymphocytes besides dendritic cells and natural killer (NK) cells playing an important role in the pathogenesis and control of viral infections [7, 8]. Lymphopenia has been recognized as a predictor of worse outcomes among COVID-19 patients [9]. Changes in phenotypes and numbers of lymphocyte subset have been reported in COVID-19 patients [10-14]. Emerging of studies showed a decrease in the frequencies of regulatory T (Tregs) cells (CD4^+^CD25^+^FOXP3^+^) and CD45RA^+^Treg cells in severe COVID-19 patients [15, 16]. The importance of the alterations of these parameters especially (T_reg_ cells), CD27^+^, and CD38^+^ lymphocytes during the disease not well documented. In the previous study we show that an increasing trend in total T cells, T helpers, cytotoxic T cells, activated lymphocytes, and natural killer cells among covid-19 patients who are responding and recovering [17].

Thus in the current study, we did details of immunophenotyping of lymphocytes at the admission of time and tried to find any correlation with clinical outcome and severity of COVID-19 disease.

## Materials and methods

### Patients

In the period of study 23 to 30 May 2020, a total of 67 confirmed COVID-19 patients were enrolled in the study at Masih Daneshvari Hospital, Tehran, Iran. The patients were selected symptomatic and the SARS-CoV-2 RNA in samples was confirmed by WHO conducted reverse transcriptase-polymerase chain reaction (RT-PCR) assay. The severity of the disease was evaluated by detecting rest oxygen saturation (O_2_sat) levels and respiratory rate. Patients with pulmonary infiltration as evaluated by chest X ray (CXR) or computerized tomography (CT) and O_2_ sat >93% with ambient air were classified as moderate groups (24 cases) and patients with O_2_ sat ≤93% or a respiratory rate of more than 30 breaths/min was categorized as severe cases (32 cases). Intensive care unit (ICU) admitted patients; the patients, who need noninvasive or mechanical ventilation; and the patients with acute respiratory syndrome distress (ARDS) or shock, were classified as critical cases (11 cases) [18, 19]. The study protocol was approved by the Ethics Committee of the national research institute of tuberculosis and lung diseases (approval number: IR.SBMU.NRITLD.REC.1399.037). Written informed consent was obtained from all participants or their legal guardians. All experiments were performed in accordance with relevant guidelines and regulations.

### Therapeutic approach

All patients were under supportive care consisting of intravenous fluid and supplemental oxygen. As the recommendation of the Iranian national guideline of COVID-19 management at the time of the study (details mentioned elsewhere [20], all patients have received lopinavir/ritonavir 400/100 mg for seven days.

### Blood sampling

Whole blood (2 ml), peripheral blood was collected in EDTA anticoagulated tubes on the first day of admission, before treatment initiation. All blood samples were tested up to six hours after sampling. Total blood counts (CBC) and CD3^+^, CD4^+^, CD8^+^ T cells; CD19^+^ and CD20^+^ B lymphocytes; CD16^+^ CD56^+^ (NK cells), and regulatory T cells (CD4/CD25 and FOXP3+) frequency and absolutes counts (cells/μl) were measured.

Briefly, the samples were centrifuged, and then red blood cells were removed by adding lysis buffer. After that white blood cells were harvested, and then washed with cold PBS. Thereafter, the cell-surface Fc receptors were blocked with 2.4 G2 (PharMingen, San Diego, CA, USA) before staining. Antibodies which is used for flow cytometry were phycoerythrin (PE)-conjugated anti-human CD4, CD19, CD16/56 antibodies (PharMingen, USA) to staining CD4 T cells, CD19 for B cells, CD16/56 for NK cells, respectively. CD8 cells were stained by anti -human CD8 conjugated with allophycocyanin (APC) antibody. For CD3 T cells, fluorescein sothiocyanate (FITC)-conjugated anti-human antibody (PharMingen, USA) was used. All antibodies were used for cells and incubated at 4 for 30 min at a dark place. To determine the immunophenotyping of Treg cells, surface staining of CD4 and CD25 markers was performed by mouse anti-human CD25-FITC (Biolegend, San Diego, CA, USA), and CD4-PE (Immunostep, Salamanca, Spain) for 30 min at 4 °C. After that, the cells were washed and then incubated with fixation and permeabilization solution buffer (BD Biosciences, San Diego, CA, USA) for 15 min at 4 °C. Thereafter, cells were washed with cold PBS and intracellular staining performed by FOXP3-APC Abs (eBioscience, California, USA) for 30 min in 4 °. Isotype-matched antibodies were used with all the samples. After that, the cells were washed and 10000 events were analyzed by FACS (FACSCalibour, USA). The data was analyzed by flowjo software version 8.

### Statistical analysis

Categorical variables were described as number and frequency, and continuous variables as median and interquartile range. Categorical data were compared using the chi-square test or Fisher’s Exact test. The Kolmogorov-Smirnov and Shapiro-Wilk tests were used to test the normality of data. For comparison of medians between different groups of severity, the Kruskal-Wallis test was used and differences between two groups were analyzed using the Mann – Whitney test. SPSS Statistics version 21.0 software was used for statistical analyses. All reported P values are two-sided and a value of less than 0.05 was considered statistically significant. The diagram was drawn using R software.

## Results

### Baseline characteristics

In the period of the study, 67 patients with COVID-19 were enrolled. The mean age of cases was 51.3 ± 14.8 years, 31 (46.3%) were male and 52.2% had minimally one comorbidity. Twenty four (35.8%) patients classified as moderate cases, 32 patients (47.8%) were classified as severe cases and 11 (16.4%) patients were categorized as critical. Demographics, basic characteristics, and outcome of treatment of patients were summarized in **table 1**. Patients with O_2_sat less than 93 were significantly older than others (54±15 versus 46±14 years, p=0.040). Also rate of underline disease (40.3% versus 11.9%, p=0.021) particularly diabetics (22.4% versus 4.5%, p=0.047) was significantly higher among hypoxic cases. Concerning outcome, mortality was more common among cases with hypertension (6%) in comparison to others (4.5%) p=0.040 **(Table 1)**. Median of age, erythrocyte sedimentation rate (ESR), and C-reactive protein (CRP) were not significantly different among patients with different severity of the disease. In the patients with a critical condition, the median of lactate dehydrogenase (LDH) was higher than severe and moderate cases (p-value 0.005 and 0.001 respectively).

**Table 1.**
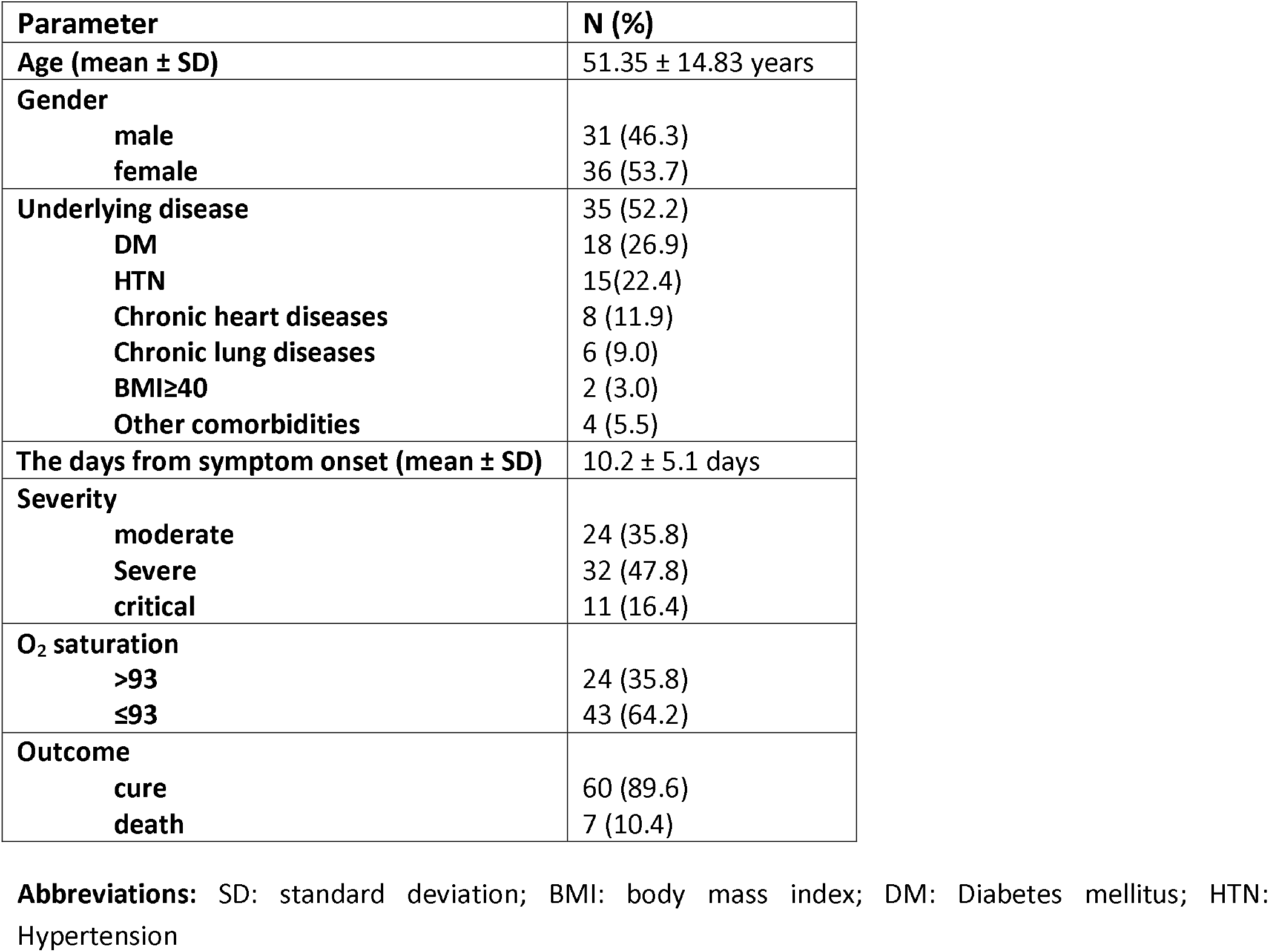
Demographic and characteristics of 67 confirmed COVID-19 cases

### Immunophenotypying and severity of the disease

Leukocyte count, percentage of neutrophils and lymphocytes, and percentage and absolute count of lymphocyte subsets were compared among patients with different severity of the disease. Cases with more severe disease had a higher percentage of blood neutrophils. Conversely, the percentage of blood lymphocytes, CD3^+^ cells, CD25^+^FOXP3^+^ T cells; and absolute count of CD3^+^ T cells, CD25^+^FOXP3^+^ T cells, CD4^+^ T cells, CD8^+^ T cells, CD16^+^56^+^ lymphocytes were lower in more severe cases in comparison to milder cases (Fig.1). No significant difference was found in other parameters as well as the CD4^+^/CD8^+^ ratio.

### Cellular subsets in moderate, severe and critical COVID-19 patients

The frequency of polymorphonuclears in critical patients were significantly higher than those in severe (p=0.0038) and moderate (p=0.00094) COVID-19 patients (p=0.00051)). Moreover, the percentage of PMNs in severe patients were higher than those in moderate patients (p=0.027) (**Fig. 1A**).The frequency of lymphocytes in critical patients were significantly lower than those in severe (p=0.0049) and moderate (p=0.0011) COVID-19 patients (p=0.00026), in addition to, the percentage of in severe patients were lower than in moderate COVID-19 patients (p=0.0065) (**Fig. 1B**). The frequency of total T lymphocytes in critical patients were not significantly difference from severe (p=0.18) and moderate (p=0.055) patients (p=0.087) **(Fig. 1C)**. The total lymphocyte count in critical patients was significantly lower than in severe (p=0.011) and moderate (p=0.00041) patients (p=0.00029). In spite of that the total lymphocyte count in severe patients had no significant difference with moderate patients (P=0.16) (**Fig. 1D**). There was significant difference at CD4+Tcell count between critical COVID-19 patients and severe (p=0.0017) and moderate group patients (p=0.00033) (**Fig. 1E**).

**Fig. 1.**
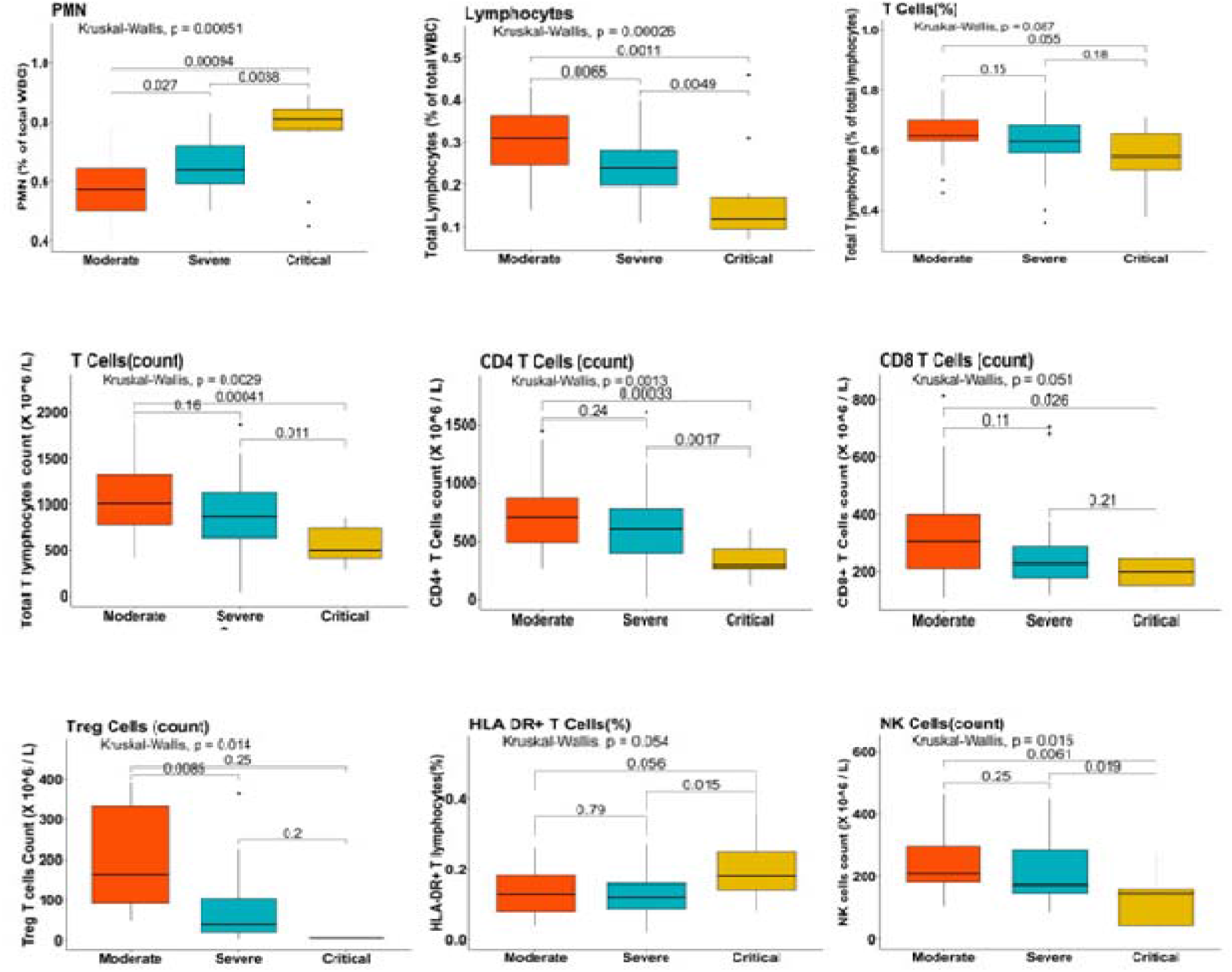
Comparison of on admission cellular subsets in COVID-19 patients with different disease severity **Abbreviations:** T_reg_: regulatory T lymphocyte (CD3^+^CD25^+^FOXP3^+^), NK: natural killer

There was no significant difference at CD8+Tcell count between critical and severe COVID-19 patients (p=0.21), on the contrary, CD8+Tcell count in critical patients was significantly higher than in moderate COVID-19 patients (p=0.026) (**Fig. 1F**).

The Treg cells count in severe COVID-19 patients was significantly lower than in moderate patients (p=0.014). However, Treg count in critical patients was similar to severe (p=0.2) and moderate (0.25) patients (**Fig. 1G**).

The frequency of HLA-DR+T lymphocytes in critical patients were significantly higher than in severe (p= 0.015) patients, but there was no significant difference in the frequency of HLA-DR+T lymphocytes in critical and moderate COVID-19 patients (p=0.056) (**Fig. 1H**). There was significant difference at NK cell frequency between critical patients and severe (p=0.019) and moderate (p=0.0061) patients (p=0.015) (**Fig. 1I**). Patients who died had a total leukocyte [median; 11380, interquartile range (IQR), 6370-15350 (cell/μl)] and neutrophil percentage in peripheral blood 78% [IQR; 73-88%] which were significantly higher than those [5775, IQR; 4140-7800 (cell/μl) and 62%, IQR; 53-72%, respectively] in cure patients (p=0.023 and p=0.001 respectively) **(Table 2)**. As well as, the percentage of lymphocytes in dead patients [12%, IQR; 8-20%] was significantly lower than cure patients [25%, IQR; 19-34%, p=0.002] **(Table 2)**. The frequency of T cells (CD3+) [60%, IQR; 38-62%] in death group was lower than in cure patients [64%, IQR; 59-69%, p=0.042)]. The NK Cell percentage [6%, IQR; 5-13%] and count [76 (cell/μl), IQR; 43-185 (cell/μl)] in death group was significantly lower than in cure group [16%, IQR; 11-18% and 204 (cell/μl), IQR; 149-300 (cell/μl), respectively. (p=0.006 and p=0.04) (**Table 2**)

**Table 2:**
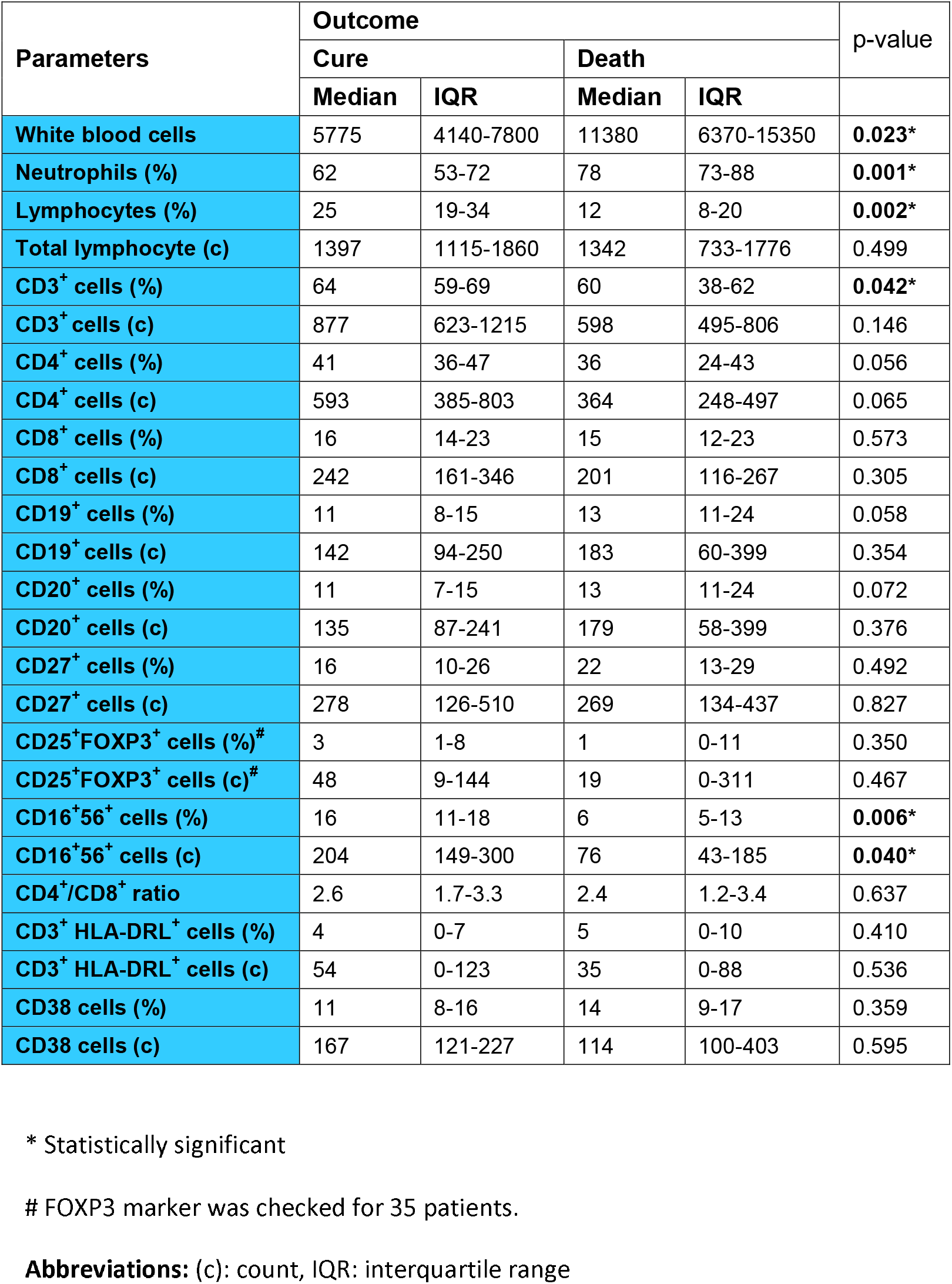
Distribution of cellular subsets among COVID-19 patients concerning outcome

| Parameters | Outcome |  |  |  | p-value |
| --- | --- | --- | --- | --- | --- |
|  | Cure |  | Death |  |  |
|  | Median | IQR | Median | IQR |  |
| White blood cells | 5775 | 4140-7800 | 11380 | 6370-15350 | **0.023\*** |
| Neutrophils (%) | 62 | 53-72 | 78 | 73-88 | **0.001\*** |
| Lymphocytes (%) | 25 | 19-34 | 12 | 8-20 | **0.002\*** |
| Total lymphocyte (c) | 1397 | 1115-1860 | 1342 | 733-1776 | 0.499 |
| CD3<sup>+</sup> cells (%) | 64 | 59-69 | 60 | 38-62 | **0.042\*** |
| CD3<sup>+</sup> cells (c) | 877 | 623-1215 | 598 | 495-806 | 0.146 |
| CD4<sup>+</sup> cells (%) | 41 | 36-47 | 36 | 24-43 | 0.056 |
| CD4<sup>+</sup> cells (c) | 593 | 385-803 | 364 | 248-497 | 0.065 |
| CD8<sup>+</sup> cells (%) | 16 | 14-23 | 15 | 12-23 | 0.573 |
| CD8<sup>+</sup> cells (c) | 242 | 161-346 | 201 | 116-267 | 0.305 |
| CD19<sup>+</sup> cells (%) | 11 | 8-15 | 13 | 11-24 | 0.058 |
| CD19<sup>+</sup> cells (c) | 142 | 94-250 | 183 | 60-399 | 0.354 |
| CD20<sup>+</sup> cells (%) | 11 | 7-15 | 13 | 11-24 | 0.072 |
| CD20<sup>+</sup> cells (c) | 135 | 87-241 | 179 | 58-399 | 0.376 |
| CD27<sup>+</sup> cells (%) | 16 | 10-26 | 22 | 13-29 | 0.492 |
| CD27<sup>+</sup> cells (c) | 278 | 126-510 | 269 | 134-437 | 0.827 |
| CD25<sup>+</sup>FOXP3<sup>+</sup> cells (%)<sup>#</sup> | 3 | 1-8 | 1 | 0-11 | 0.350 |
| CD25<sup>+</sup>FOXP3<sup>+</sup> cells (c)<sup>#</sup> | 48 | 9-144 | 19 | 0-311 | 0.467 |
| CD16<sup>+</sup>56<sup>+</sup> cells (%) | 16 | 11-18 | 6 | 5-13 | **0.006\*** |
| CD16<sup>+</sup>56<sup>+</sup> cells (c) | 204 | 149-300 | 76 | 43-185 | **0.040\*** |
| CD4<sup>+</sup>/CD8<sup>+</sup> ratio | 2.6 | 1.7-3.3 | 2.4 | 1.2-3.4 | 0.637 |
| CD3<sup>+</sup> HLA-DRL<sup>+</sup> cells (%) | 4 | 0-7 | 5 | 0-10 | 0.410 |
| CD3<sup>+</sup> HLA-DRL<sup>+</sup> cells (c) | 54 | 0-123 | 35 | 0-88 | 0.536 |
| CD38 cells (%) | 11 | 8-16 | 14 | 9-17 | 0.359 |
| CD38 cells (c) | 167 | 121-227 | 114 | 100-403 | 0.595 |

\* Statistically significant

## Discussion

In this study, we found a significant differences in the counts of total lymphocytes and immunophenotyping of lymphocytes (CD4^+^ cells, CD8^+^ cells, CD25^+^FOXP3^+^ regulatory T cells, NK cells), in the severe disease. In the pathogenesis of COVID-19 disease immune dysregulation induced by SARS-CoV-2 has been shown [21, 22]. In this regard, the virus may cause immune system exhaustion [5, 8], cytokine storm, and hyper inflammation [23-25]. Lymphocyte and their subsets have a great role in the adequacy of the immune system response against viral infections [7] and any insult to this population induces dysregulation of the immune system [10, 13, 26]. Lymphopenia and decrease of CD4^+^ and CD8^+^ subsets of T cells were reported in the cases of the SARS-CoV [27]. Several studies show a significant decrease in the total number of lymphocytes among COVID-19 patients [21, 28, 29].

Moreover. emerging of studies in COVID-19 patients reported a correlation between cellular subsets with the disease severity and but not much information available in linked to the outcome of disease [10, 13, 15, 21, 30, 31]. A strong association was found between low levels of total lymphocytes, CD4^+^ T cells, CD8^+^ T cells, and the disease severity in COVID-19 patients [8,13-15, 21, 26].

In this regard, CD4^+^ T cells play an important role in the regulation of immune response against viral infections. This function is performed by activation of the network between immune cells via secretion of pro and anti-inflammatory mediators. Besides this function, some subsets of T cells can lyse the tumor cells and infected cells. In this line, CD8^+^ cytotoxic T cells are able to kill host cells infected by the virus [32]. The exact mechanism of lymphopenia and reduction of the counts of T cells in COVID-19 disease is not yet elucidated. Few studies emphasize SARS-CoV-2 directly able to damage lymphocytes, especially T cells [15]. Chu et al. demonstrated that MERS-CoV (another Betacoronavirus) could able to infect and induce apoptosis iof T cells, however, this was not observed with the SARS virus infection [33] however speculated that play a role in the pathogenesis of COVID-19 disease. Recent study shows infiltration and sequestration of lymphocytes mainly CD8^+^ T cells in affected organs, especially pulmonary interstitium [29]. In SARS-CoV-infected patients, it is showed that about 80% of total infiltrative inflammatory cells in the lungs are CD8^+^ T cells [34]. Shift of the immune cells from blood to the tissue as shown in some autopsies [29, 35] may explain the lower count of circulating T cells, especially CD8^+^ cells in severe COVID-19 patients.

Weiskopf et al. showed an increase in CD4:CD8 ratio in ten moderate to severe COVID-19 patients [36]. Moreover, Diao et al. reported this ratio between patients under ICU care, and other admitted cases found a higher proportion among the first group [21]. However, in the current study we did not show a significant difference in this ratio between severe and non-severe COVID-19 patients [14, 15].

The reason for this finding may be a parallel reduction of both T cell subsets in severe cases. Regulatory T cells (T_reg_) are essential components for immune homeostasis and inhibit exaggerated inflammatory responses following viral infections [37]. Qin et al showed lower blood levels of T_reg_ cells among COVID-19 patients especially in severe cases [15]. In the current study, a correlation between T_regs_ and the severity of the disease was found. However, in study performed by Chen et al. despite the reduction of T_regs_ among both severe and moderate cases, the total Treg cells was comparable between the two groups [26]. They show that in severe cases a significantly lower proportion of naive T_regs_ and a slightly higher proportion of memory T_regs_. As T_reg_ lymphocytes able to inhibit the overreaction of the immune response to pathogens, their reduction may accounts for the tissue damage in severe cases of COVID-19 patients [38].

B cells frequency and total in COVID-19 patients was inconsistent. In this line, Chen et al. reported the count of B lymphocytes not significantly different concerning the severity of the disease [26]. However, they found a higher proportion of B cells in severe cases and accounted for somewhat related to a more significant decrease in T cells in this group of patients. Conversely, Wang et al and in a meta-analysis, a significant reduction of B cells among more severe cases was shown. The mechanism of this process is not clearly defined [10, 14].

Although studies did not find any correlation between the counts of NK cells and severity of COVID-19 disease [14, 26], the current study data in line with a meta-analysis performed by Huang [10] who is reported lower levels of NK cells among severe COVID-19 patients [39].

In our study, a higher number of polymorphonuclear cells; and a lower proportion of lymphocytes, T cells, and NK cells were shown which is suggestive as a predictors of mortality of COVID-19 patients.

For supporting of this hypothesis the presented data shows that in patients with normal T cells lower ration of mortality than with patients with lower T cells counts.In correspondence, recently two studies have shown a correlation of CD4^+^ and CD8^+^ T cell counts with the death of COVID-19 patients [14, 21].

In summary the current study has limitations due to the variability and diverse future of COVID-19 which unable to generalize the statement. Concerning the complexity of the immune system, prospective multicenter studies with a large sample of patients need to be studied. Besides, analysis of cellular subsets in detail with multi-colored staining of subsets of T cells such as Th9, Th17 and Th22 T cells are necessary to assess the time-based changes in immune response after infection with SARS-Co-V-2 and their prognostic value.

In conclusion, our data suggests that the counts of total lymphocyte and their immunophenotyping (consisting of CD4 ^+^, CD8^+^, CD25^+^FOXP3^+^ T cells, and NK cells) may be correlated with the severity of COVID-19 patients. The reduction of these cells may suggests for usage on them as the predictors of the severity of the disease, however, need to be explored in detail in the future.

## Data Availability

Not applicable

## Statements

## Acknowledgement

The authors acknowledge from all participants and clinicians and nurses for helping and reminding the memory all patients whom died from COVID-19 during this study.

## Statement of Ethics

All protocol has been approved by ethic committee of Maish Daneshvari Hospital.

## Conflict of Interest Statement

There is no conflict interest in this work.

## Funding Sources

Not applicable

## Author Contributions

MR, MM, NHK and EM wrote first draft. NKD, ND and EM did experiments. HRJ, PT,AM, DM,ZA,MP provided the patients and characherization. MKD did statistical analysis. MV and AAV approved final version.

## Notes

### Competing Interest Statement

The authors have declared no competing interest.

### Author Declarations

The study protocol was approved by the Ethics Committee of the national research institute of tuberculosis and lung diseases (approval number: IR.SBMU.NRITLD.REC.1399.037).

